# High-resolution advanced diffusion MRI of rectal cancer surgical specimens: correlating microstructural characteristics with histology

**DOI:** 10.64898/2026.04.02.26350055

**Authors:** Ana R Fouto, Hasti Calá, Shermann Moreira, Noam Shemesh, Laura Fernandez, Nuno Couto, Ignacio Herrando, Stephanie Nougaret, Raluca Popita, Jorge Brito, Susana Ouro, Miguel Chambel, Nikolaos Papanikolaou, Amjad Parvaiz, Richard J. Heald, Mireia Castillo-Martin, Inês Santiago, Andrada Ianus

## Abstract

**Background:** Despite advances in organ-preserving strategies for rectal cancer, accurate restaging after neoadjuvant therapy (NAT) remains challenging due to the limited sensitivity of conventional MRI in differentiating residual tumour from treatment-induced changes. This limitation highlights the urgent need to develop better imaging tools that can accurately analyze the complex structure of the treated rectal wall.

**Purpose:** To study the diffusion properties of different rectal wall components, including healthy layers and pathological tissue, using high-resolution ex vivo diffusion MRI (dMRI) on whole total mesorectal excision (TME) samples obtained after NAT, and to evaluate how advanced diffusion metrics improve tissue analysis compared to standard T_2_-weighted imaging.

**Materials and Methods:** Five post-NAT TME specimens were prospectively collected at a single center and fixed (36h formalin, 4h PBS). Then, specimens were mounted in Fomblin and scanned using a 9.4T Bruker BioSpec (22°; 86 mm Tx/Rx). Diffusion MRI was acquired using a 2D multi-shell sequence (TR/TE 11,000/24 ms; 130 slices; 0.5 mm³ isotropic voxel; b = 1500 and 3000 s/mm²; 15 directions) alongside multi-echo T_2_-weighted imaging (TR 25,000 ms; 8 echoes; TE 10–80 ms; fat suppression). Diffusion and kurtosis parametric maps were generated by voxelwise linear least-squares fitting; T_2_ maps by monoexponential fitting (MATLAB). Specimens were sectioned at 5 mm, stained with H&E and dual staining (for fibrosis and smooth muscle), digitized, and co-registered with MRI using morphological landmarks. Regions-of-interest (ROIs) - mucosa, submucosa, muscle layers, tumour, and fibrous tissue - were compared using a linear mixed-effects model with FDR correction (RStudio v2025.09).

**Results:** The *muscularis propria* exhibited the highest FA values of all tissue components, reflecting the ordered fiber architecture of its inner circular and outer longitudinal layers, which were visually separable on direction-encoded colour FA maps. Focal disruption of anisotropy at the tumour-muscle interface corresponded histologically to tumour invasion of the *muscularis propria*. Tumour regions showed the lowest mean diffusivity (MD), reflecting high cellularity and restricted diffusion, and MD was comparatively higher in the residual scar. Kurtosis metrics - particularly MK and AK - were elevated in tumour, reflecting greater microstructural heterogeneity and complexity. T_2_ mapping provided limited contrast across tissue types due to formalin fixation effects.

**Conclusion:** Diffusion MRI metrics quantitatively discriminated rectal wall tissue components ex vivo with histological validation, beyond T_2_-weighted contrast. DTI and DKI metrics characterized tumour, fibrous tissue, and *muscularis propria* invasion, supporting their potential as microstructural imaging biomarkers for treatment response assessment.

## 1. Introduction

Rectal cancer is a major and growing global health concern (1,2), and accurate local restaging and treatment response evaluation after neoadjuvant therapy (NAT) are crucial for optimal clinical management (3,4). A critical decision for patients with locally advanced rectal cancer (LARC) is whether to pursue surgical resection or organ-preserving strategies such as Watch-and-Wait (5,6). This critical decision relies on pelvic MRI to differentiate treatment-induced fibrosis and edema from residual tumour following neoadjuvant therapy (NAT), as well as to assess the depth of tumour infiltration, thereby guiding personalized treatment strategies (7,8).

The standard imaging technique for both initial staging and post-treatment evaluation in rectal cancer is T_2_-weighted imaging. However, its ability to distinguish between fibrosis and residual tumour is limited (9,10). To overcome this limitation, there is increasing interest in diffusion MRI (dMRI) as an adjunct to conventional imaging (4). The benefit of employing dMRI is its ability to provide information about the tissue’s microstructure and cellular density (11). Currently, in clinical settings, the persistent hyperintensity on the high b-value diffusion-weighted images reflects restricted water diffusion and is the most common clinical proxy for identifying malignant tissue (12–14).

Moreover, additional quantitative information can also be derived from diffusion data. It was previously shown that the high apparent diffusion coefficient (ADC) values result from water molecules diffusing more freely, which happens when the treatment is successful, whereas lower ADC values usually indicate higher cellularity and residual tumour (12,15). However, diffusion in biological tissues is often directionally dependent due to organized structural components such as smooth muscle fibers and stromal architecture, which can impact the direct interpretation of ADC values. Therefore, the use of advanced diffusion imaging techniques such as diffusion tensor imaging (DTI) for a more comprehensive analysis is often needed (16). DTI extends conventional ADC analysis by modeling anisotropic diffusion and enabling the estimation of parameters such as fractional anisotropy (FA) and mean diffusivity (MD) (17,18). In the rectal wall, anisotropy may reflect the layered organization of muscular structures, while tumour infiltration can disrupt this directional coherence. Despite its potential, DTI has been applied to rectal cancer in only a small number of studies, and the biological interpretation of its parameters in the context of rectal wall microstructure requires histological validation (12,14,19). Moreover, other advanced diffusion techniques, such as diffusional kurtosis imaging (DKI), can further account for deviations from Gaussian diffusion and provide metrics that are more sensitive to the complexities of tissue microstructure (20). Kurtosis parameters have been increasingly explored in clinical studies, particularly for differentiating residual tumour from fibrosis after NAT and for improving response assessment (21–24). To our knowledge, however, no prior study has applied DKI to ex vivo rectal cancer specimens, representing a gap in the microstructural characterization of rectal wall tissue components. Thus, DKI can potentially provide complementary information to ADC and DTI for characterizing tissue microstructure.

To establish the biological validity of imaging biomarkers, it is important to understand the link between MRI-derived metrics and underlying tissue microstructure. Therefore, imaging of surgical specimens enables high-resolution acquisition and direct correlation with histopathology. Although prior ex vivo diffusion MRI investigations in rectal cancer have suggested correlations between diffusion metrics and cellular density or stromal composition, they were limited to small tissue samples and a restricted set of quantitative parameters - namely FA, ADC, and T_2_ contrast - without non-Gaussian diffusion modeling or comprehensive histologically validated characterization of individual rectal wall layers (12,15).

In this study, we performed detailed ex vivo high-resolution quantitative MRI of whole total mesorectal excision (TME) specimens obtained after NAT to investigate, for the first time, advanced diffusion and T_2_ properties of distinct components of the rectal wall and pathological changes. We aimed to determine whether diffusion and relaxometry metrics can reliably distinguish tumour, fibrosis, and individual wall layers. Thereby providing a stronger microstructural basis for MRI-based treatment response assessment and more informed decision-making in rectal cancer management.

## 2. Materials and Methods

All experiments were approved by the institutional ethics committee.

### 2.1. Patients and specimen preparation for ex-vivo MRI

The study processed specimens from six patients with rectal cancer who had received neoadjuvant chemoradiotherapy (nCRT) at the Digestive Unit of the Champalimaud Clinical Centre. All patients had signed a consent to allow the use of their biological specimens for research purposes, and all samples were pseudonymized by the Champalimaud Foundation Biobank. After surgery, the fresh TME surgical specimen was sent to the Service of Anatomic Pathology with a cold ischemia time of less than 20 minutes. After checking the condition of the mesorectum, the front and back surfaces that are not covered by the peritoneum were marked with blue and black ink, respectively. Then, the specimen was cut open lengthwise starting from the top along the front side until it reached the peritoneal reflection and put in 10% buffered formalin for 36 hours at room temperature (RT), with gauze inside the rectal lumen to help with the fixing process. After fixation, the specimen was thoroughly rinsed in running water for 15 minutes and later placed for 4 h in 1xPBS at RT. The sample was mounted in a cylindrical container filled with Fomblin and used for imaging.

### 2.2. MRI data acquisition

TME specimens preserved in the containers were then scanned using a 9.4T Bruker Biospec system at 22°C (86 mm transmit/receive). Figure 1 illustrates a schematic of specimen preparation and MRI data acquisition workflow. Subsequently, a high-resolution MRI imaging protocol was used to evaluate the microstructural properties of the rectal tissue. It included a 2D standard diffusion-weighted imaging and a 2D multi-slice multi-echo (MSME) that showed standard T_2_-weighted contrast.

**Figure 1.**
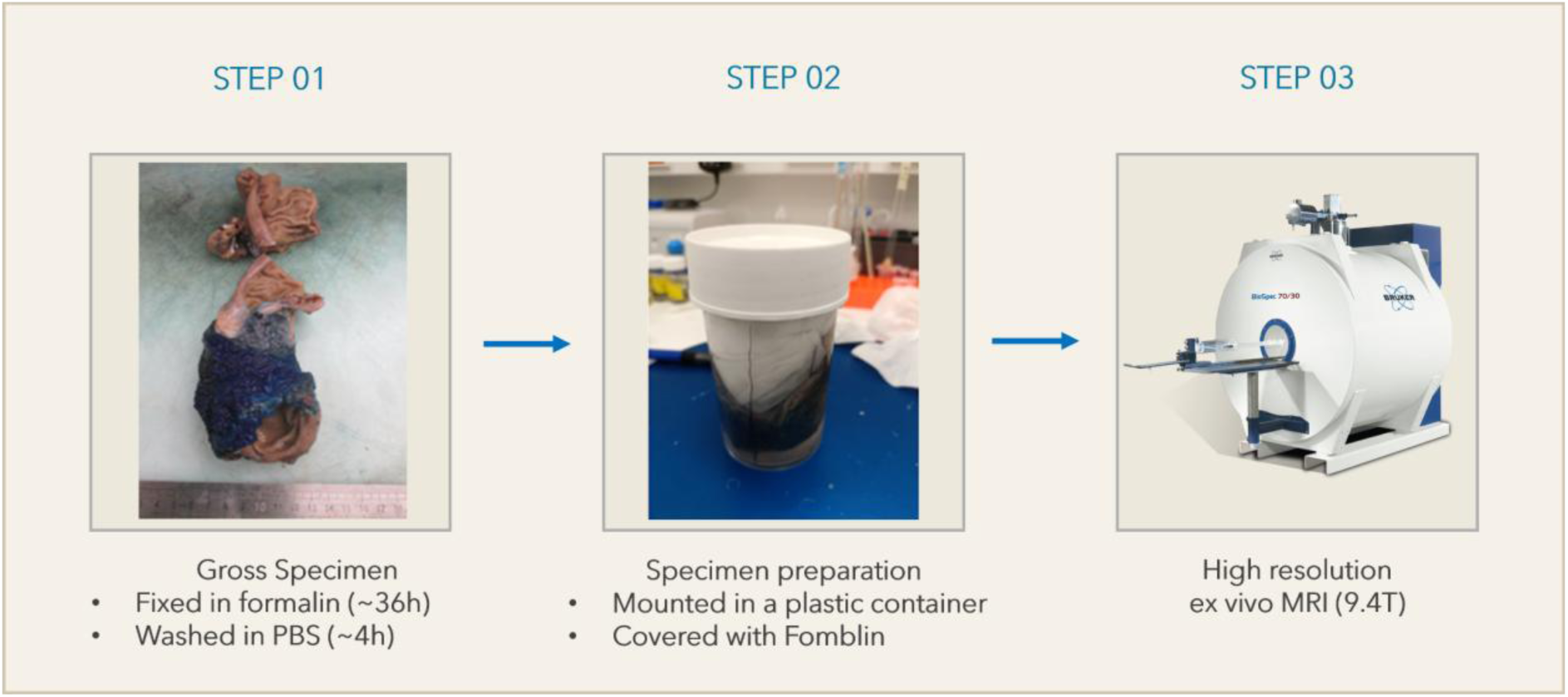
- Schematic overview of ex vivo specimen preparation and MRI acquisition workflow. STEP 01 **-** After the surgery, blue ink was used to mark the front of the fresh total mesorectal excision (TME) specimen, and black ink was used to mark the back. The specimen was then cut open along the front side up to the peritoneal reflection and fixed in 10% buffered formalin for about 36 hours at room temperature, with gauze put inside the rectum to help with the fixation. STEP 02 - After being fixed, the specimen was washed in PBS for four hours at room temperature, put in a cylindrical container filled with Fomblin, and scanned with a Bruker Biospec 9.4T MRI system (STEP 03).

dMRI data were collected with the following parameters: TR/TE=11000/24 ms; 130 slices; matrix size: 140 x 130; isotropic resolution of 0.5 mm³, 2 b0 images, 2 b-values (1500 s/mm², 3000 s/mm²); and 15 diffusion directions. The MSME sequence was defined with TR = 25000 ms, eight echoes (TEs ranging from 10 to 80 ms), and the same geometric parameters. We applied fat suppression techniques to both sequences.

The high-resolution MRI images from the TME sample, which included S_0_ (showing standard T_2_-weighted contrast) and the average signal at s/mm², are shown in Figure 2 to demonstrate the overall appearance and quality of the data.

**Figure 2.**
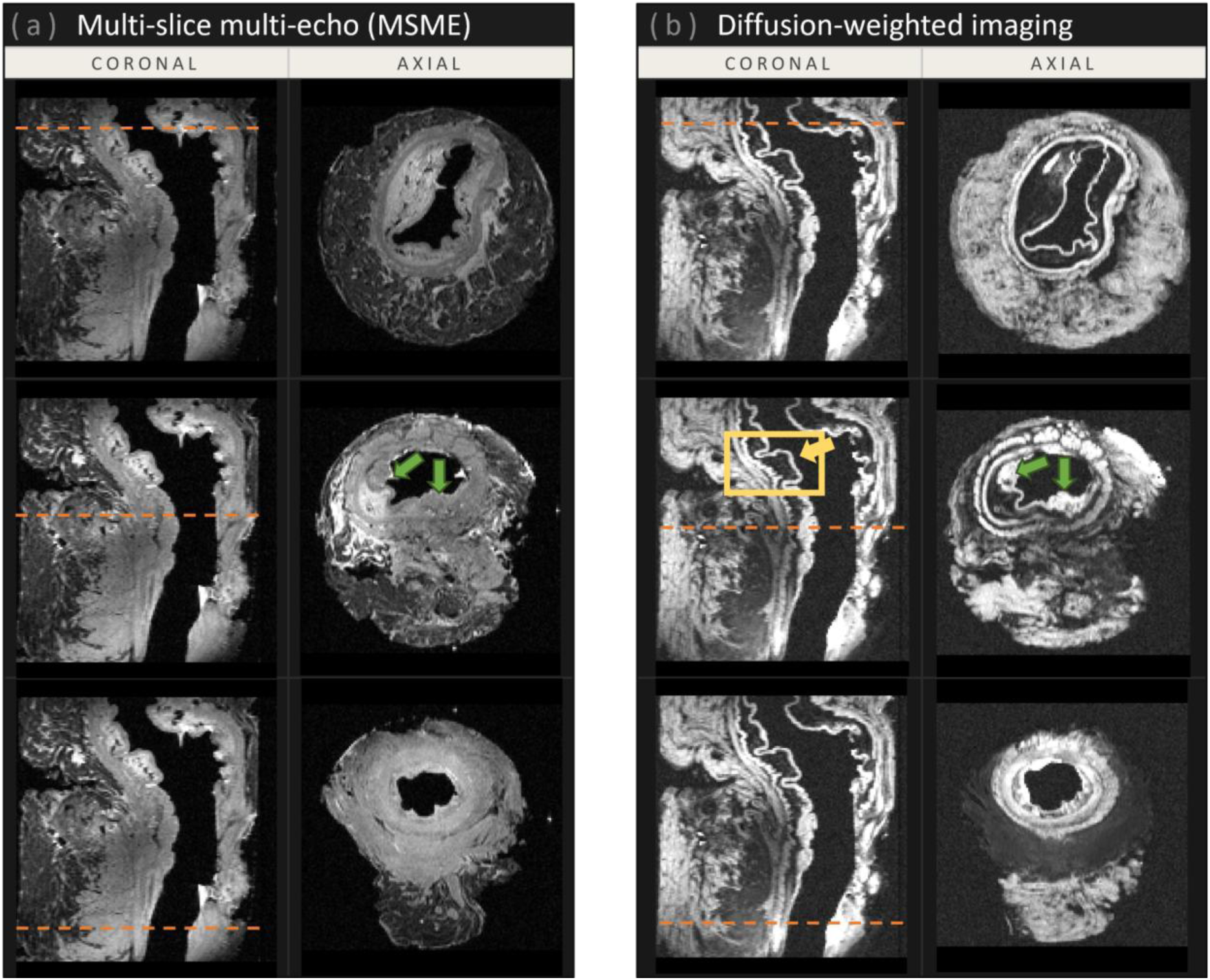
- Representative images of the imaging protocol collected from the ex vivo specimen. Examples of (a) multi-slice multi-echo (MSME), which shows T_2_-weighted imaging contrast, and (b) the average signal at b=3000 s/mm^2^. Both contrasts primarily highlight structural boundaries between tissue types (yellow box), with diffusion-weighted images providing greater detail than T_2_-weighted images. Tumour regions (indicated by green arrows) are also more clearly delineated in the average signal at b=3000 s/mm^2^ compared to standard T_2_ contrast. Dashed orange lines indicate the corresponding anatomical slice position.

### 2.3. Histopathology examination

The specimen was cut transversally into 5 mm sections beginning at the distal end to obtain a series of cross sections, and selected areas were used for further processing to generate paraffin-embedded tissue blocks, which were cut into 5 μm-thick sections and stained with standard hematoxylin and eosin (H&E).

*Dual modified trichromic and smooth muscle actin (sma) immunohistochemistry (IHC) staining:* To better characterize the fibrosis and differentiate it from residual smooth muscle bundles, a dual staining was developed by sequentially performing a standard chromogenic immunohistochemistry for sma followed by a modified Masson’s trichrome staining following previous reports (25).

Briefly, after the 5 µm sections were deparaffinized and rehydrated, they were rinsed in 1X Phosphate Saline Buffer (PBS) (Mediatech, Corning) and incubated in 1% H_2_O_2_ (8.3 mL H_2_O_2_ up to 250 mL 1X PBS) for 15 minutes at RT. Antigen retrieval buffer (pH 9) was performed in a steamer for 30 minutes. After the slides cooled down, they were incubated with blocking solutions (2% Bovine Serum Albumin (BSA) in 1X PBS) for 30 minutes at RT. Then, the slides were incubated with the primary antibody against alpha-smooth muscle actin diluted in blocking serum (Abcam, ab7817, 1:200), for 2 hours at RT followed by incubation with HRP-conjugated anti-mouse secondary antibody (ThermoFisher, cat#31432, 1:200) in blocking solution for 1 hour at RT. Immunoreactivity was visualized using 3,3’-Diaminobenzidine (DAB) chromogen (Thermo Scientific, H54000.06) in 0.5% Triton X-100 (Fisher Bioreagents, BP151-500) solution for 10 minutes. The reaction was stopped by immersing the slides in dH_2_O, and staining intensity was checked with a brightfield microscope.

Following immunohistochemistry staining, the slides were subjected to a modified Masson’s Trichrome using the Trichrome Stain Kit (Connective Tissue Stain) (Abcam, ab150686). Briefly, Bouin’s fluid was preheated to 65°C in a water bath under a fume hood, and slides were immersed in the preheated solution for 60 minutes, followed by a 10-minute cooling period at RT. They were then rinsed thoroughly in running tap water until completely clear, followed by staining with Weigert’s iron hematoxylin, previously obtained by mixing equal parts of Weigert’s solution A and B, for 5 minutes, followed by rinsing under running tap water for 2 minutes. Aniline Blue solution was used for 1 minute and subsequently rinsed in dH_2_O. After that, the slides were put into a 1% acetic acid solution for 5 minutes. Finally, the slides were dehydrated, cleared in xylene, and mounted using a xylene-based mounting media (Leica Biosystems, cat#3801733).

All histological slides were digitized using a Philips Ultra-Fast Scanner 1.6, thumbnail images were obtained, and they were manually reconstructed (Figure 3). They were then matched to MRI slices using grossing images as reference (see section 2.4.2).

**Figure 3.**
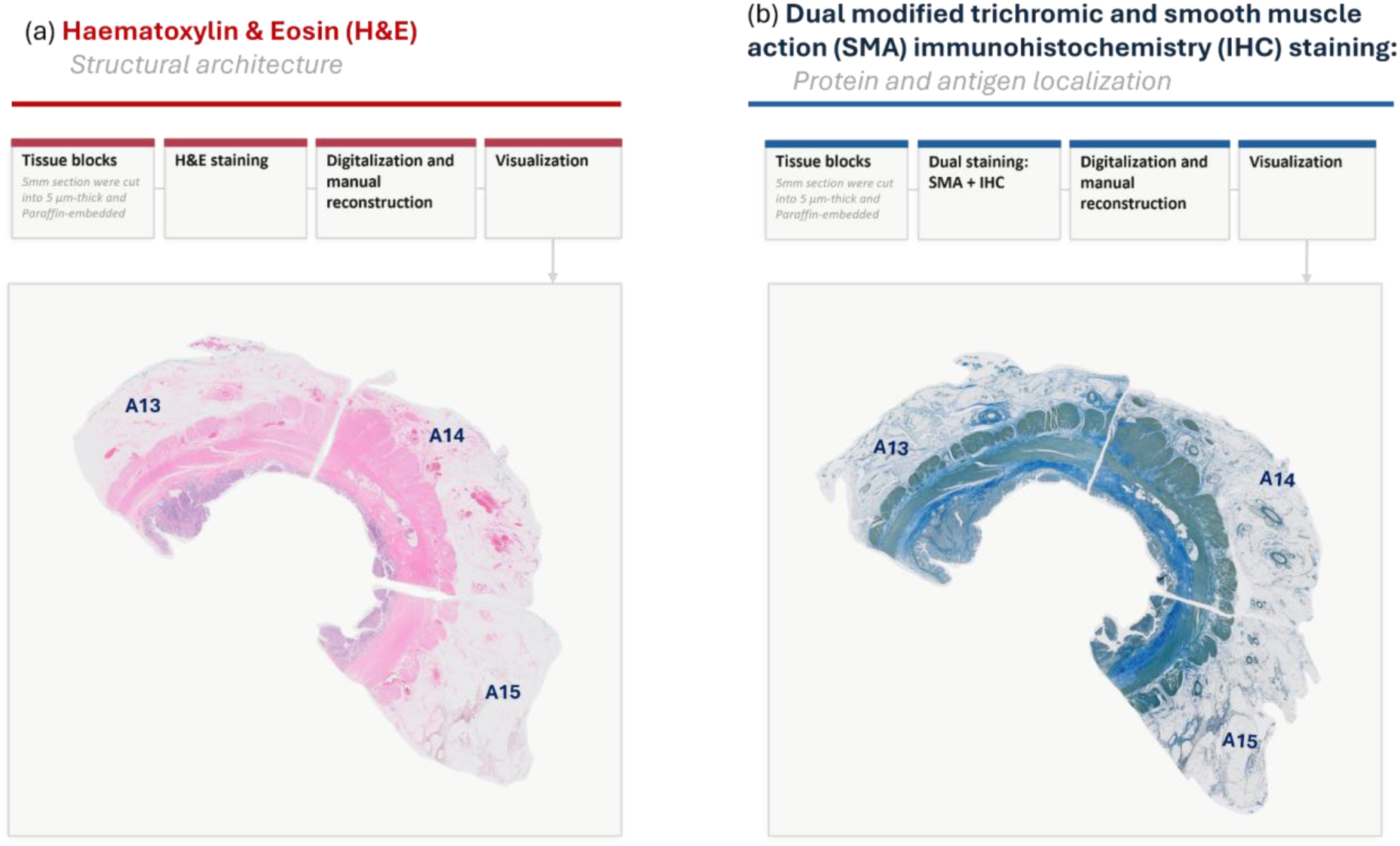
- Illustrative examples of the histological staining analysis: **(a)** standard haematoxylin and eosin (H&E); and **(b)** dual staining combining smooth muscle actin (SMA) immunohistochemistry (IHC) and modified Masson’s trichrome (right), used to differentiate fibrosis (blue) from residual smooth muscle bundles (brown).

### 2.4. MRI data analysis

#### 2.4.1. MRI data preprocessing and parametric maps extraction

Data conversion and subsequent MRI analyses were performed using a custom MATLAB-based pipeline. All MRI datasets were visually inspected for artifacts and inconsistencies, and one specimen was excluded from further analysis due to fat artifacts.

Initially, we limited the quantitative analysis to the rectal wall tissue by generating a binary mask from the first echo of the multi-echo T_2_ data. We used percentile-based intensity thresholding, followed by morphological filtering to remove small, isolated regions and the background.

Prior to fitting the model, the diffusion data were cleaned up using complex-valued denoising. Next, we used a voxelwise linear least squares (LLS) algorithm to estimate the diffusion and kurtosis tensor parameters. Lastly, we extracted maps of fractional anisotropy (FA), mean diffusivity (MD), axial diffusivity (AD), radial diffusivity (RD), mean kurtosis (MK), axial kurtosis (AK), and radial kurtosis (RK). We also calculated the T_2_ maps from the multi-echo data by fitting a simple exponential model to each voxel across eight different echo times (all generated maps were saved in the NIfTI format). Figure 4 depicts an illustrative example of the appearance of the maps obtained.

**Figure 4.**
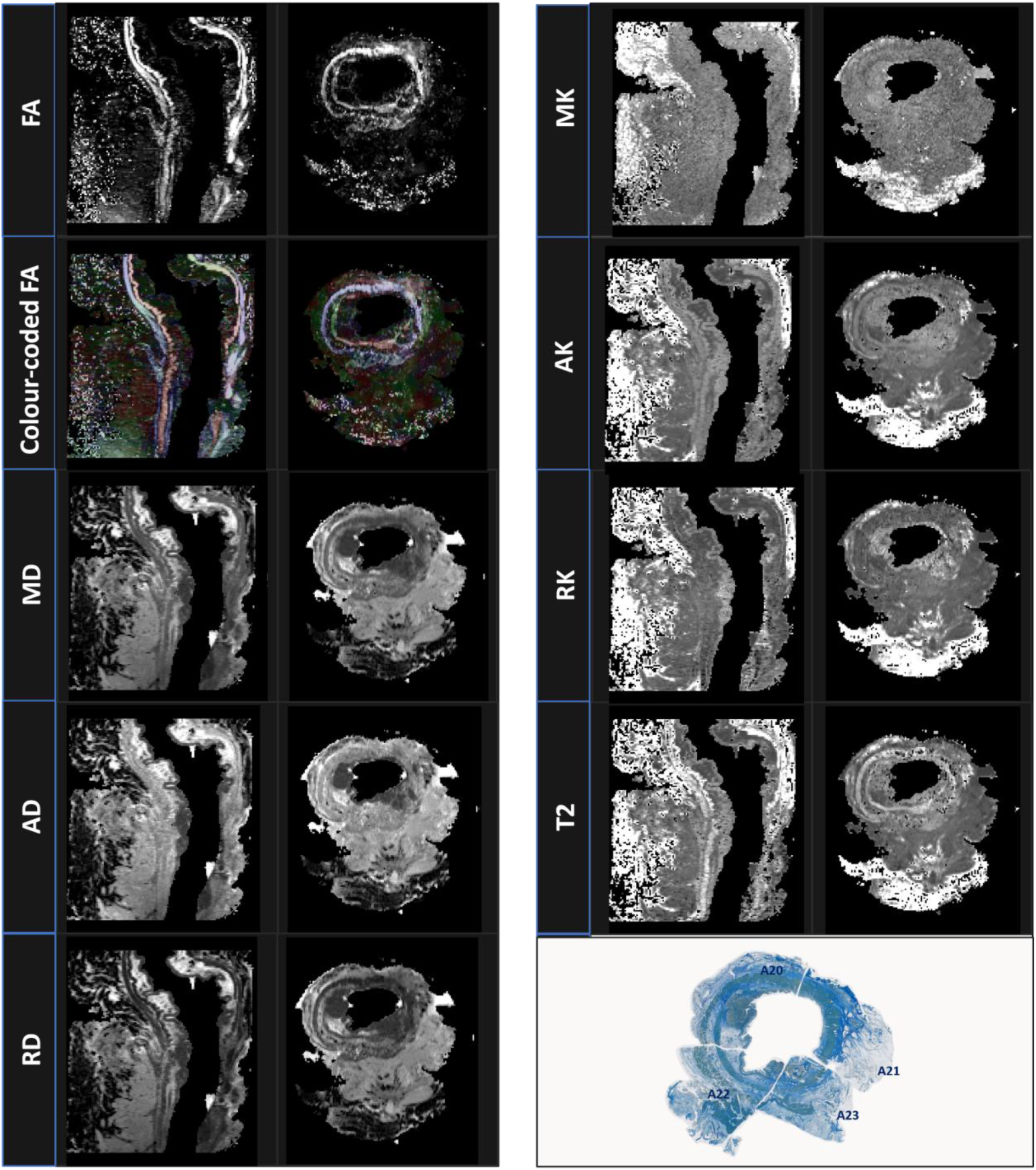
- Illustrative slices of the quantitative MRI-derived maps of the MRI data obtained from the ex vivo specimen: (1) Fractional Anisotropy (FA); (2) Mean diffusivity (MD; in mm^2^/s); (3) Axial diffusivity (AD; in mm^2^/s); (4) Radial diffusivity (RD; in mm^2^/s); (5) Mean kurtosis (MK); (6) Axial kurtosis (AK); (7) Radial kurtosis (RK); (8) T_2_ map (ms); and (9) the corresponding manually reconstructed axial histology slice stained with dual staining that distinguishes fibrosis (blue) from residual smooth muscle bundles (brown).

#### 2.4.2. MRI-histopathology-guided regions-of-interest (ROI) analysis

MRI-histopathology correlation was performed to guide the manual definition of anatomically and biologically informed regions of interest (ROIs) of the rectal wall components: mucosa (M); submucosa (S); circular muscle layer (CL); longitudinal muscle layer (LM); tumour (T); and fibrous tissue (FT). This procedure was carried out collaboratively by a pathologist with 24 years of experience and a radiologist with 14 years of experience.

Firstly, the matching MRI slices were iteratively selected for each specimen by visually identifying shared morphological landmarks (e.g., wall thickness variations, lumen configuration, tumour size and shape, etc.) on the digitized H&E histology slices as well as the overall specimen shape provided by grossing images. After reviewing the FA, MD, and high-b-value averaged images, the team chose the MRI slice that had the closest morphological correspondence to a given histology slice.

Following slice matching, annotated distinct tissue components on the histology images were used as biological reference points for tumour, muscle layers, mucosa, submucosa, and fibrotic regions.

The ROIs were placed to correspond spatially with the histology annotations, which were then manually defined using FSLeyes. When we were unsure about the H&E staining or MRI features, particularly in distinguishing fibrotic tissue, we did extra histological staining (see section 2.3) to better understand it. As a result, all ROIs were reviewed and refined based on the updated histopathological information. Figure 5 depicts the step-by-step workflow for defining rad-path. Figure 6 illustrates ROI drawing after slice selection. Finally, a qualitative MRI-histopathology assessment was performed to investigate the ability of diffusion MRI contrasts to visually identify tumour infiltration patterns within the rectal wall.

**Figure 5.**
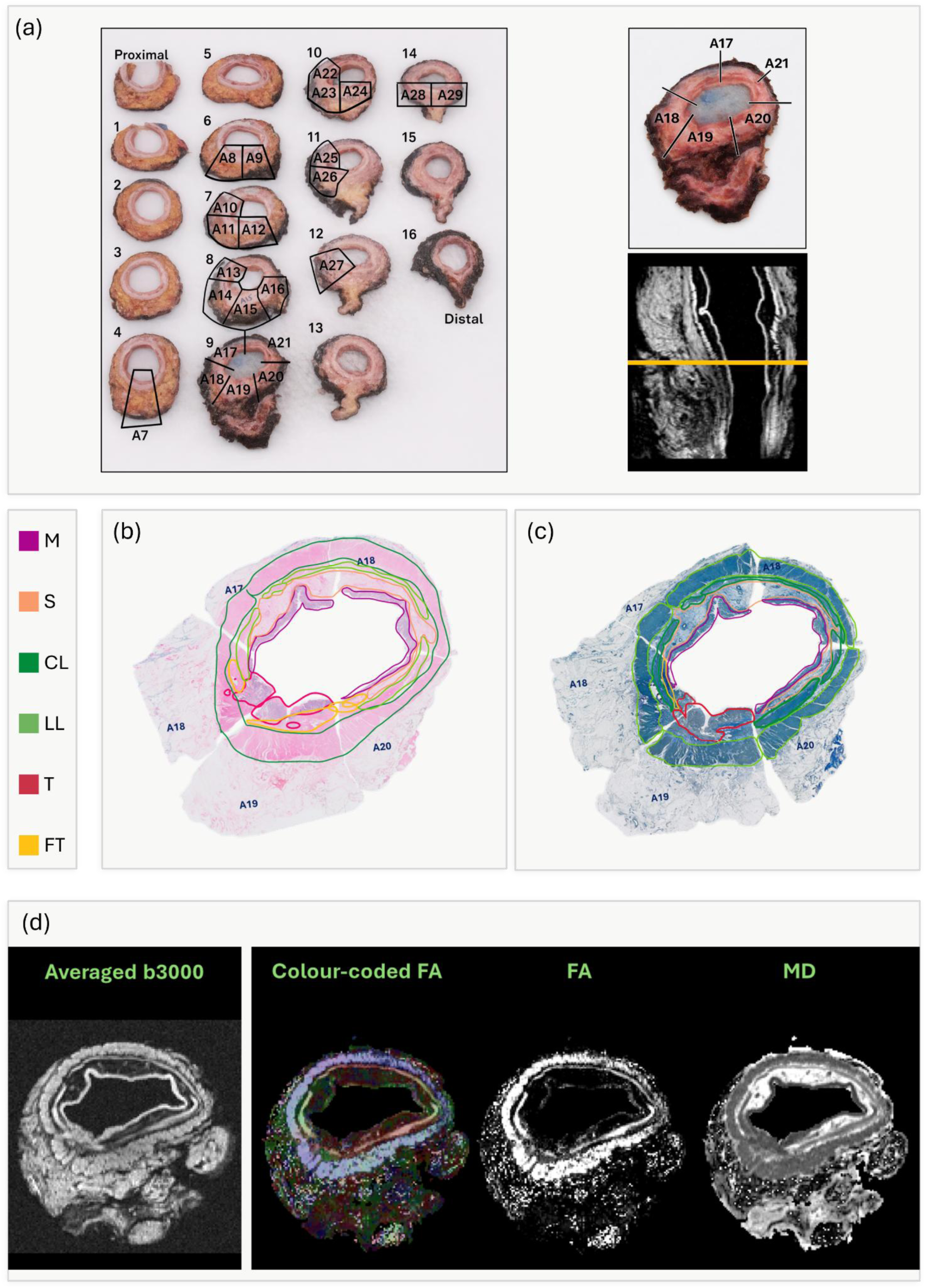
- Workflow for matching histology slices with diffusion MRI data. Grossing images (a) are first used to approximate the location of the corresponding MRI slice on the averaged b = 3000 s/mm² diffusion-weighted image. After digital reconstruction of the histology slices, histological landmarks visible on the stained sections (b) and (c), such as tumors or muscles, are then used to refine the slice selection. The final match is made by lining up the anatomical landmarks (d) on all the diffusion-weighted images and diffusion MRI parameter maps. M - Mucosa; S - Submucosa; CL - Circular muscle layer; LL - Longitudinal muscle layer; T - Tumour; FT - Fibrous Tissue.

**Figure 6.**
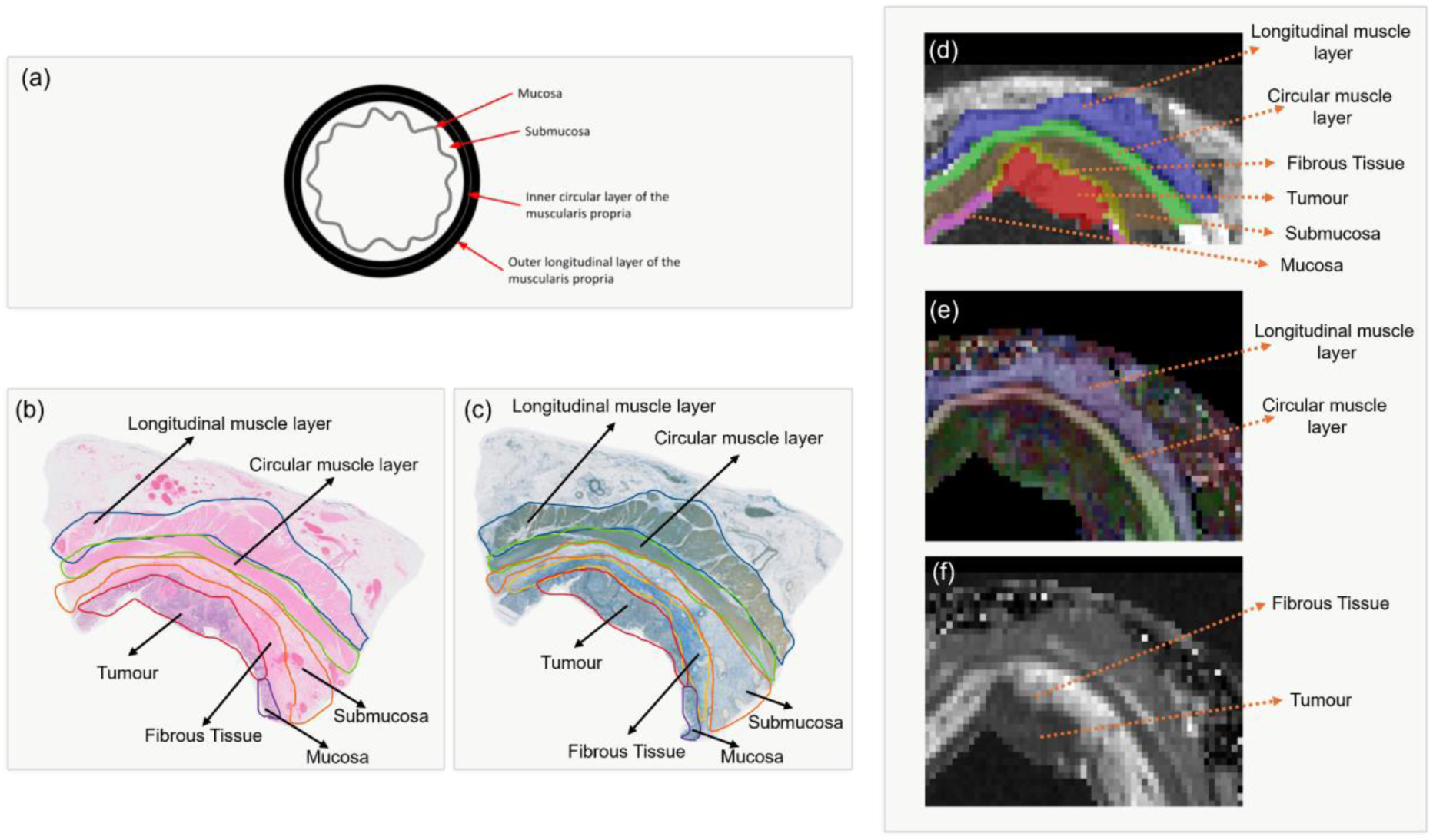
- Illustrative example of the workflow used for region-of-interest (ROI) delineation based on histology-MRI correspondence. (a) Schematic representation of the healthy rectal wall layers. (b) Standard hematoxylin and eosin (H&E) staining and (c) dual staining combining smooth muscle actin (SMA) immunohistochemistry and modified Masson’s trichrome used to identify anatomical structures. After slide preparation and digital reconstruction, an experienced pathologist annotated and validated key anatomical landmarks on the histology sections (b,c). These landmarks were then matched to the corresponding MRI slice through rad-path correlation and validated by a radiologist. Colour-coded fractional anisotropy (FA) maps (e) and mean diffusivity (MD) maps (f) were used together with the averaged b = 3000 s/mm² diffusion-weighted image (d) to guide ROI placement and ensure anatomical correspondence with the histological structures. Finally, ROIs (d) were manually delineated to represent specific tissue types: mucosa (purple), submucosa (copper), longitudinal muscle layer (dark blue), circular muscle layer (green), tumour tissue (red), and fibrous tissue (yellow). All masks shown in (d) are displayed with transparency.

### 2.5. Statistical analysis

Median values were employed to summarize quantitative MRI parameters for each ROI using custom Python scripts. Next, we used a linear mixed-effects model with FDR correction for multiple comparisons to assess differences in median values for each parametric map (i.e., diffusion parameter and T_2_ map) across six tissue types. The tissue type was considered a fixed effect, and the subject was included as a random effect. Post-hoc pairwise comparisons were performed with Tukey adjustment. To further manage the issue of comparing multiple tissue types and maps, the Tukey-adjusted p-values were also adjusted using the Benjamini–Hochberg false discovery rate (FDR Statistical analyses were conducted using RStudio (v-2025.09.2+418), and P < 0.05 was considered statistically significant.

## 3. Results

### 3.1. Qualitative MRI-histopathology findings

Comparing histopathology and MRI showed a link between the biology of the tissue and the signal observed in diffusion imaging. The *muscularis propria* showed a weaker signal on the high b-value diffusion-weighted image, while the healthy mucosa showed a strong bright signal. Nonetheless, the tumour areas were also of bright intensity, contrasting with the surrounding muscle layers’ weaker signal, resulting in a visually detectable tissue contrast. However, its anatomical distribution was different and extended beyond the normal mucosal surface, even though the healthy mucosa signal was equally bright. In summary, visual examination of MRI-histopathology-matched slices revealed two distinct patterns of pathological tissue signal change within the rectal wall: (1) direct invasion of the *muscularis propria* and (2) discrete, small tumour *foci* separate from the primary lesion.

#### Invasion of the muscularis propria

Tumor infiltration into the muscular layer revealed a distinct pattern on the diffusion maps, as shown in Figure 7. The FA maps show the spread of adenocarcinoma cells in the muscle layer very clearly. The disturbance of the normal muscle structure in these areas results in a lower FA compared to the surrounding healthy muscle. Moreover, the size and location of these changes closely matched the size of the tumor observed in the tissue samples.

**Figure 7.**
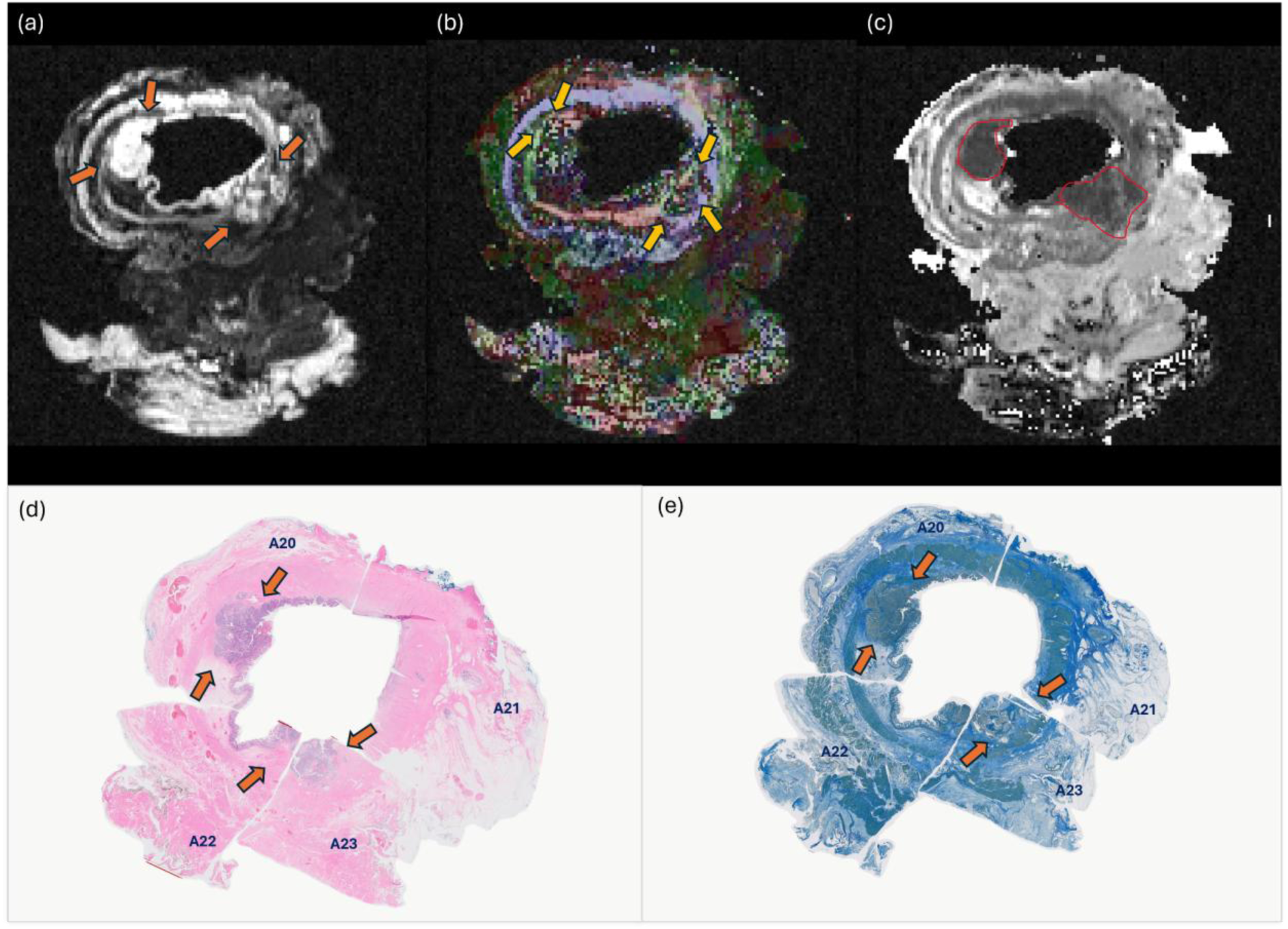
- Comparison of MRI contrasts and histology illustrating tumour invasion in rectal muscle layers. (a) Averaged diffusion-weighted image at b = 3000 s/mm², (b) Colour-coded fractional anisotropy (FA) map, and (c) mean diffusivity (MD) map are shown alongside corresponding histology (d) H&E and (e) dual staining combining SMA IHC and modified Masson’s trichrome. FA maps clearly reveal disruptions in muscle tissue (yellow arrows), which align with areas of adenocarcinoma infiltration identified on histology and averaged b3000 (orange arrows) and MD map (red contour).

### 3.2. Differentiation of rectal wall tissue components using parametric maps

The measurements from the healthy rectal wall and the tumor areas showed clear patterns that likely indicate changes in the types of cells and their arrangement, which are important for better clinical re-staging evaluations. In summary, FA showed the organization of the muscular layers, MD highlighted tumor-related diffusion restriction, and kurtosis offered supplementary details on the microstructural heterogeneity of tumour regions compared to surrounding tissues. Figures 8 and 9 illustrate the distributions of median values and the corresponding between-tissue differences across parametric maps, highlighting significant contrasts across the different components of rectal wall tissue.

**Figure 8.**
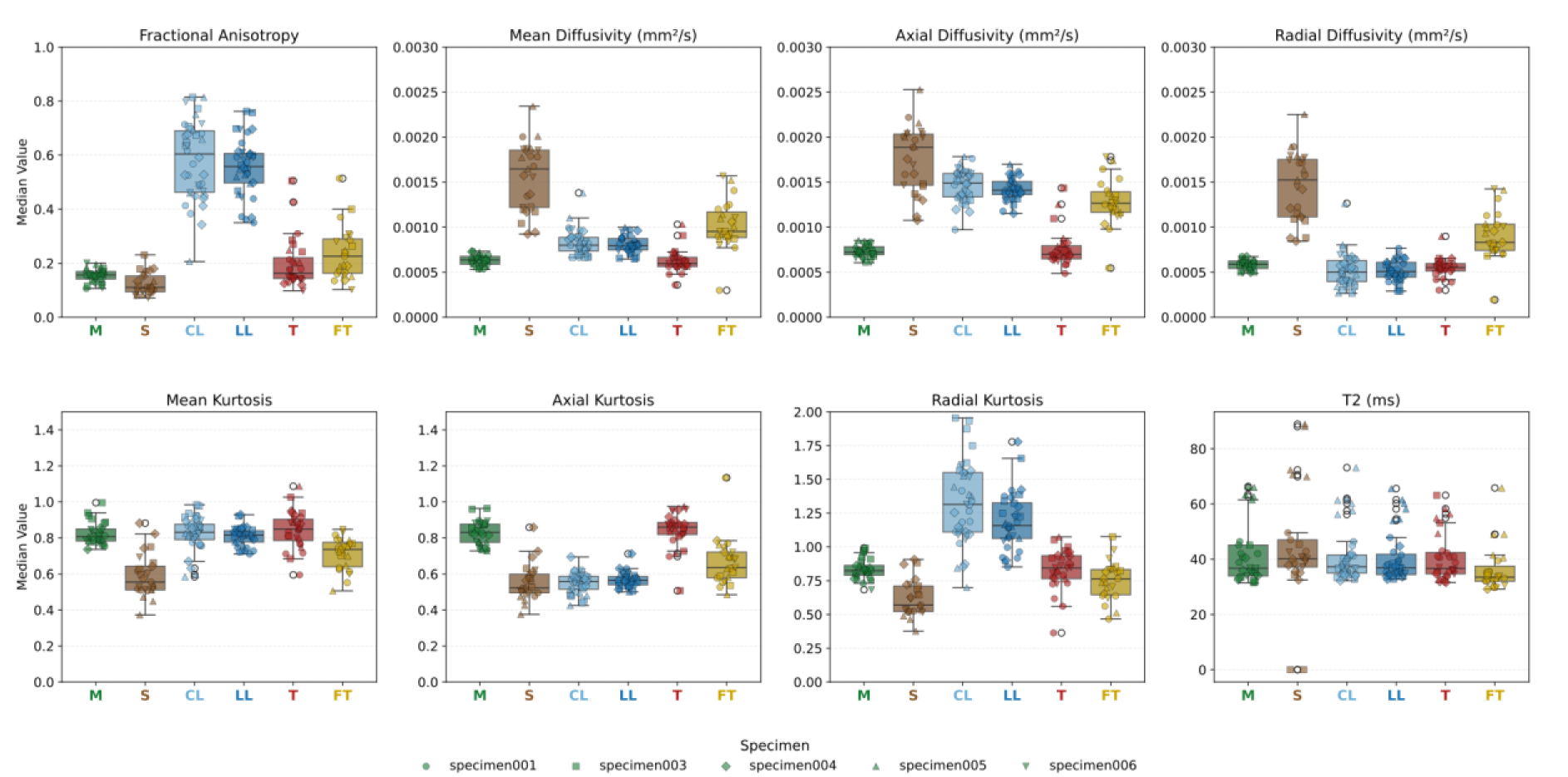
- Boxplots depict the distribution of quantitative diffusion MRI metrics across the normal layers of the rectal wall (mucosa (M), submucosa (S), circular muscle layer (CL), and longitudinal muscle layer (LL)) and pathological tissue (tumour (T) and fibrous tissue (FT)). The metrics: fractional anisotropy (FA), mean diffusivity (MD), axial diffusivity (AD), radial diffusivity (RD), mean kurtosis (MK), axial kurtosis (AK), radial kurtosis (RK) - show distinct diffusion patterns for healthy and pathological tissues.

**Figure 9.**
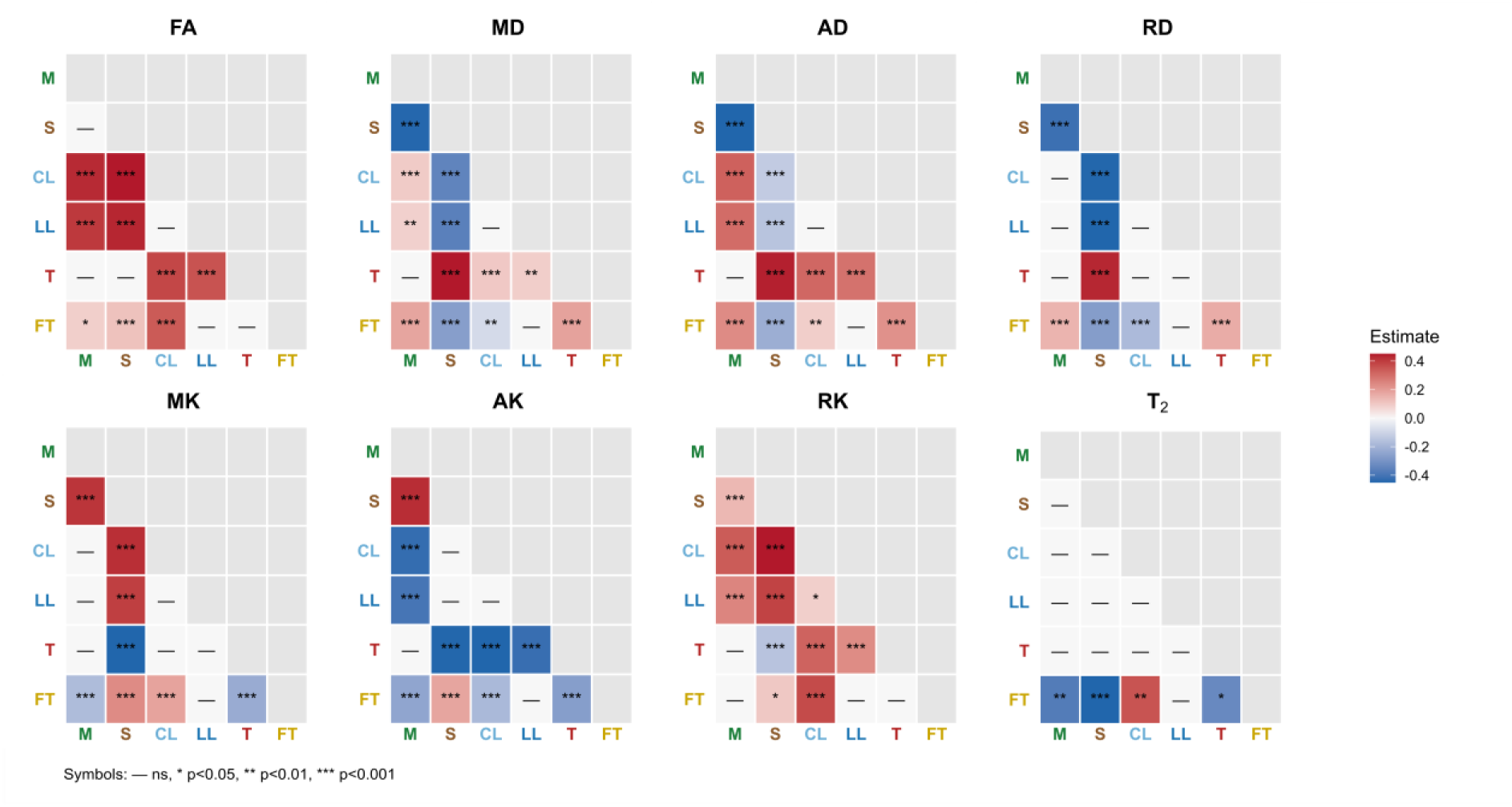
- Matrix summarizing significant differences in diffusion MRI metrics (fractional anisotropy (FA), mean diffusivity (MD), axial diffusivity (AD), radial diffusivity (RD), mean kurtosis (MK), axial kurtosis (AK), and radial kurtosis (RK)) across rectal wall tissues - mucosa (M), submucosa (S), circular muscle layer (CL), longitudinal muscle layer (LL) - and pathological tissues - tumour (T) and fibrous tissue (FT). Tumour and fibrous regions exhibit distinctive diffusion patterns relative to healthy tissue layers, reflecting underlying microstructural alterations relevant for tissue characterization and tumour detection. *** P<0.001; ** P<0.01; * P<0.05. The colour scale indicates the magnitude of the statistical estimates.

#### 3.2.1. Diffusion MRI parameters

##### Normal rectal wall layers

FA values clearly showed an anisotropic pattern across the normal layers of the rectal wall. The *muscularis propria* exhibited the highest FA values, while the mucosa and submucosa had lower values. Furthermore, the circular layer displayed greater dispersion. Importantly, FA clearly differentiates between the circular and longitudinal muscle layers and other non-muscle tissues, with minimal overlap with other types of tissue. Direction-encoded FA maps revealed clear alignment of principal diffusion orientation (V1) and distinct separation between the inner circular and outer longitudinal muscle layers. Within the normal rectal wall, MD also demonstrated a layer-dependent diffusivity pattern. The mucosa had the lowest MD values, while the submucosa had higher and more variable values. The *muscularis propria* exhibited intermediate MD values, characterized by partial overlap between the longitudinal and circular layers. Within the muscle layers, AD values remained steady in the main direction of diffusion, whereas RD was lower, indicating restricted diffusion perpendicular to the fiber orientation. In the layers of the inner rectal wall, we also found that, similar to MD, AD and RD were lower in the mucosa and higher in the submucosa.

Kurtosis metrics varied depending on compartment-specific patterns across the rectal wall layers. MK was higher in the muscle layers and mucosa and lower in the submucosa. AK was elevated in the mucosa and lower in both the submucosa and muscle layers, while RK was higher in the muscle layers.

##### Pathological tissue comparisons Tumour vs muscularis

At the tumour-muscle interface, both the tumour and *muscularis propria* restrict diffusion. However, FA and V1 orientation clearly distinguish tumour infiltration from intact muscle, as *muscularis propria* retains high anisotropy and ordered fiber direction, while infiltrated regions show reduced FA and lower MD. This enables precise identification of tumour extension into muscle.

##### Tumour vs fibrous tissue (residual tumour vs scar)

Diffusion maps showed that both tumour and fibrous tissue had similar patterns of anisotropy (FA), and both had higher FA than mucosa. However, FA was not a reliable discriminator between tumour and fibrous tissue due to significant overlap. In contrast, MD was lowest in tumour regions, while fibrous tissue displayed slightly higher MD values. AD and RD were also lower in tumours in comparison to fibrotic tissue, with a more pronounced reduction in RD, indicating greater transverse diffusion restriction. Kurtosis metrics, particularly MK and AK, were higher in tumours than in fibrous tissue, reflecting greater microstructural heterogeneity than the surrounding tissues. Fibrous tissue generally exhibited lower kurtosis values.

#### 3.2.2. T_2_ relaxation characteristics

T_2_ mapping demonstrated limited inter-compartment discrimination, yielding insufficient contrast for tissue-type differentiation compared with the diffusion-derived parametric maps. Mucosa and submucosa had slightly higher T_2_ values, but distinctions between tumour, fibrous tissue, and deeper wall layers were much clearer on diffusion and kurtosis maps. Moreover, tumour demonstrated slightly higher T_2_ relaxation times compared with fibrous tissue.

## 4. Discussion

Advanced diffusion MRI at 9.4T, including diffusion tensor and kurtosis imaging, quantitatively characterized distinct rectal wall tissue compartments and pathological tissue in *ex vivo* specimens, with all findings histologically validated. We found tissue-specific diffusion patterns that underline clinical imaging observations. FA mapped the structural organization of the *muscularis propria*, reflecting the ordered fiber architecture of its smooth muscle layers, while MD differentiated tumour from fibrous tissue through its sensitivity to cellularity-driven diffusion restriction. Kurtosis metrics captured microstructural heterogeneity within tumour, consistent with the non-Gaussian diffusion behavior expected in architecturally complex tissue. T_2_ mapping, in contrast, showed limited contrast across tissues. Together, these results support the potential of advanced dMRI as a quantitative biomarker framework for tumour characterization and treatment response assessment in rectal cancer.

### Muscularis propria integrity and tumour mural invasion

The inner circular and outer longitudinal layers of the *muscularis propria* exhibited the highest FA values across all tissue types. This is consistent with the highly ordered parallel fiber architecture of smooth muscle. FA measures the directionality of water diffusion, which, in organized structures, diffuses preferentially along the fiber axis, yielding elevated FA (16,18). The corresponding diffusion profile of higher AD and lower RD confirmed preferential water movement along the aligned muscle fibers. Notably, while AK was low - consistent with structural coherence along the principal fiber axis - elevated MK and RK indicated greater microstructural complexity in directions perpendicular to the main fiber orientation, likely reflecting the contribution of cross-fiber compartments to non-Gaussian diffusion behavior. This interpretation is supported by prior ex vivo DTI work in rectal cancer at ultra-high fields. Pham et al. also demonstrated at 11.7T that *muscularis propria* exhibits the highest FA values relative to non-muscular rectal wall layers and pathological tissue (12). Our results at 9.4T reinforce that FA is a sensitive and specific marker of muscular rectal wall structural integrity.

Direction-encoded colour FA maps visually separated the inner circular from the outer longitudinal muscle layer through their orthogonal fiber orientations, corroborating and extending the observations of Pham et al. at 11.7T (12). At the tumour–muscle interface, histologically validated ROIs demonstrated clear FA reduction and MD reduction in tumour compared with adjacent *muscularis propria*, consistent with the replacement of highly ordered smooth muscle architecture by the disorganized, hypercellular environment of malignant infiltration. This is directly supported by Pham et al., who demonstrated that DTI enabled detection of tumour infiltration into the *muscularis propria* through FA - identifying it as a potential novel biomarker of rectal cancer stromal organization and infiltration, particularly relevant to T1 versus T2 tumour discrimination (12).

### Tumour versus treatment-related fibrosis: the diagnostic boundary

Both the tumour and fibrous tissue exhibited higher FA than the mucosa, reflecting the structural reorganization imposed by desmoplastic collagen deposition in the tumour stroma and reactive fibrosis. However, FA alone was insufficient to discriminate between them due to substantial value overlap. MD proved more diagnostically discriminating: tumour showed the lowest MD values across all tissue types, consistent with high cellularity and restricted diffusion, while fibrous tissue demonstrated intermediate diffusivity, reflecting lower cellular density and a more open extracellular architecture despite its structural organization.

Kurtosis metrics - MK and AK - were elevated in tumour, reflecting the microstructural heterogeneity imposed by irregular cell arrangement. One recent study suggested that higher kurtosis in more invasive rectal tumours directly reflects this progressive microstructural complexity (23). Notably, fibrous tissue showed lower kurtosis than tumour despite similar FA values, suggesting that kurtosis compared to the anisotropy measures is the more sensitive discriminator of active malignancy from treatment-related fibrosis. This distinction is clinically relevant: after chemoradiotherapy, non-Gaussian diffusion behavior increases in the treated tumour bed, making kurtosis metrics particularly suited to identifying residual viable tumour (24).

### Inner wall layers discrimination: mucosa and submucosa

The mucosa demonstrated low FA and low MD, AD, and RD, reflecting the isotropic restriction imposed by its densely packed glandular epithelium, which constrains water movement uniformly and without a preferred axis. Despite this restriction, elevated MK, AK, and RK values indicate a highly compartmentalized microenvironment, consistent with the co-existence of glandular lumina, dense cytoplasm, and extracellular matrix as distinct diffusion barriers. The high signal intensity on averaged high b-value images is a practical expression of this restrictive environment and provides a reliable landmark for mucosal boundary segmentation.

On the other hand, the submucosa showed the highest diffusivity with higher MD, AD, and RD and the lowest FA and kurtosis of all rectal wall layers. Its loose connective tissue, abundant interstitial space, and low cellular density permit unrestricted, isotropic water displacement with minimal compartmentalization. Typically, the two inner layers are indistinguishable, appearing as a single intermediate-signal layer. Our results show that the submucosa’s distinct diffusivity profile sets it apart from both the mucosa above and the *muscularis propria* below, offering a quantitative basis for layer-level discrimination that conventional imaging cannot provide.

### T₂ relaxometry and the effect of ex vivo conditions

T₂ values demonstrated limited contrast across tissue types in this study, with only fibrous tissue showing slightly lower values compared with other tissue components. It is important to consider that while this finding departs from the established clinical perception that T_2_-weighted imaging provides meaningful tissue contrast in rectal cancer, it is most plausibly explained by the ex vivo experimental conditions. More specifically, formalin fixation and storage at room temperature (22°C) alter the T₂ relaxation properties of tissue, reducing free water content and shifting the equilibrium of temperature-dependent behavior of the water-protein interaction (26). These changes have a direct impact on the relaxation properties of tissue compartments, reducing the contrast that would otherwise be exploitable in living tissue at body temperature. This limitation demonstrates an inherent trade-off in ex vivo ultra-high-field studies: while spatial resolution, signal-to-noise ratio, and geometric distortion are all improved over in vivo imaging, the altered tissue state introduces quantitative biases - particularly for relaxometry-based parameters - that must be carefully considered when interpreting absolute values. In contrast, diffusion-derived parameters appear to be more resistant to these fixation-related changes, as evidenced by the present data, which show clear and histologically validated tissue discrimination using FA, MD, and kurtosis metrics.

### Advanced diffusion modelling: complementary insights across tissue compartments

DTI and DKI metrics provided complementary characterization of rectal wall microstructure. DTI metrics (i.e., FA and MD) captured tissue organization and cellularity-driven diffusion restriction, while kurtosis metrics quantified microstructural heterogeneity and complexity. Together, these parameters characterized restriction, fiber organization, and heterogeneity across tissue types in a way that neither conventional ADC nor T_2_-weighted imaging achieved in ex vivo settings or previous studies (12,15). Although absolute metric values will differ under in vivo conditions, the microstructural contrasts identified here provide a mechanistic foundation for interpreting clinical diffusion data and inform the design of advanced dMRI protocols for rectal cancer staging and treatment response assessment.

### Limitations

This study has several limitations. First, the small sample size may restrict the generalizability of our results. Second, all imaging was performed ex vivo using a 9.4T MRI scanner on formalin-fixed tissue at 22°C, which may alter tissue behavior relative to in vivo conditions and conventional clinical environments; in particular, absolute values of diffusion metrics are likely to differ from those obtained in living subjects. Third, a potential for spatial mismatch exists between the two-dimensional histological sections and the corresponding MRI slices, which could affect the precision of direct tissue-level comparisons despite the co-registration methodology employed.

## 5. Conclusion

In conclusion, this study demonstrated that using advanced high-resolution diffusion MRI metrics, especially using FA and MD, enabled the identification of the different parts of the rectal wall components and effectively distinguished healthy and pathological tissues. Diffusion-derived metrics reliably detected tumour infiltration into muscle layers, with findings closely matching histopathology results. These methods provided superior tissue characterization and anatomical detail compared to morphological T_2_-weighted imaging. Our findings support the integration of advanced diffusion MRI into clinical protocols for improved noninvasive evaluation, patient stratification, and future research in rectal cancer assessment.

## Data Availability

All data produced in the present study are available upon reasonable request to the authors.

## Acknowledgements

This work was supported by the Fundação para a Ciência e a Tecnologia (2023.02201.RESTART; THCS/0003/2023) within the framework of the co-funded Transforming Health and Care Systems (THCS) partnership (GA N° 101095654) under the EU Horizon Europe Research and Innovation Programme. AI is also supported by the ERC Horizon Europe Research and Innovation Programme Starting Grant No. 101164674.

